# Mapping rare protein-coding variants on multi-organ imaging traits

**DOI:** 10.1101/2024.11.16.24317443

**Authors:** Yijun Fan, Jie Chen, Zirui Fan, Julio Chirinos, Jason L. Stein, Patrick F. Sullivan, Rujin Wang, Ajay Nadig, David Y. Zhang, Shuai Huang, Zhiwen Jiang, Peter Yi Guan, Xinjie Qian, Ting Li, Haoyue Li, Zehui Sun, Marylyn D. Ritchie, Joan O’Brien, Walter Witschey, Daniel J. Rader, Tengfei Li, Hongtu Zhu, Bingxin Zhao

**Affiliations:** Graduate Group in Applied Mathematics and Computational Science, University of Pennsylvania, Philadelphia, PA 19104, USA; Department of Biostatistics, University of North Carolina at Chapel Hill, Chapel Hill, NC 27599, USA; Department of Statistics and Data Science, University of Pennsylvania, Philadelphia, PA 19104, USA; Division of Cardiovascular Medicine, Hospital of the University of Pennsylvania, Philadelphia, PA 19104, USA; University of Pennsylvania Perelman School of Medicine, Philadelphia, PA 19104, USA; Department of Genetics, University of North Carolina at Chapel Hill, Chapel Hill, NC 27599, USA; UNC Neuroscience Center, University of North Carolina at Chapel Hill, Chapel Hill, NC 27599, USA; Regeneron Genetics Center, 777 Old Saw Mill River Rd., Tarrytown, NY, 10591, USA; Department of Biomedical Informatics, Harvard Medical School, Boston, MA 02115, USA; Stanley Center for Psychiatric Research, Broad Institute of MIT and Harvard, Cambridge, MA 02142, USA; Program in Medical and Population Genetics, Broad Institute of MIT and Harvard, Cambridge, MA 02142, USA; Analytic and Translational Genetics Unit, Massachusetts General Hospital, Boston, MA 02114, USA; Department of Genetics, Perelman School of Medicine, University of Pennsylvania, Philadelphia, PA 19104, USA; Department of Medicine, Perelman School of Medicine, University of Pennsylvania, Philadelphia, PA 19104, USA; Institute for Biomedical Informatics, Perelman School of Medicine, University of Pennsylvania Philadelphia, PA 19104, USA; Scheie Eye Institute, University of Pennsylvania, Philadelphia, PA 19104, USA; Penn Medicine Center for Ophthalmic Genetics in Complex Diseases, Philadelphia, PA 19104, USA; Department of Radiology, Perelman School of Medicine, University of Pennsylvania, Philadelphia, PA 19104, USA; Department of Radiology, University of North Carolina at Chapel Hill, Chapel Hill, NC 27599, USA; Biomedical Research Imaging Center, School of Medicine, University of North Carolina at Chapel Hill, Chapel Hill, NC 27599, USA; Department of Computer Science, University of North Carolina at Chapel Hill, Chapel Hill, NC 27599, USA; Department of Statistics and Operations Research, University of North Carolina at Chapel Hill, Chapel Hill, NC 27599, USA; Center for AI and Data Science for Integrated Diagnostics, Perelman School of Medicine, University of Pennsylvania, Philadelphia, PA 19104, USA; Population Aging Research Center, University of Pennsylvania, Philadelphia, PA 19104, USA; Institute for Translational Medicine and Therapeutics, University of Pennsylvania, Philadelphia, PA 19104, USA; Penn Center for Eye-Brain Health, Perelman School of Medicine, University of Pennsylvania, Philadelphia, PA 19104, USA

## Abstract

Human organ structure and function are important endophenotypes for clinical outcomes. Genome-wide association studies (GWAS) have identified numerous common variants associated with phenotypes derived from magnetic resonance imaging (MRI) of the brain and body. However, the role of rare protein-coding variations affecting organ size and function is largely unknown. Here we present an exome-wide association study that evaluates 596 multi-organ MRI traits across over 50,000 individuals from the UK Biobank. We identified 107 variant-level associations and 224 gene-based burden associations (67 unique gene-trait pairs) across all MRI modalities, including *PTEN* with total brain volume, *TTN* with regional peak circumferential strain in the heart left ventricle, and *TNFRSF13B* with spleen volume. The singleton burden model and AlphaMissense annotations contributed 8 unique gene-trait pairs including the association between an approved drug target gene of *KCNA5* and brain functional activity. The identified rare coding signals elucidate some shared genetic regulation across organs, prioritize previously identified GWAS loci, and are enriched for drug targets. Overall, we demonstrate how rare variants enhance our understanding of genetic effects on human organ morphology and function and their connections to complex diseases.

---

Magnetic resonance imaging (MRI)-derived traits enable us to study the structure, function, and abnormalities of human organs *in vivo*. Many of these traits serve as established endophenotypes implicated in complex diseases and related traits. Therefore, it is of great interest to uncover genetic effects using imaging data to better understand the biology of human organs in health and disease. Recent genome-wide association studies (GWAS) have successfully identified common variants associated with multi-organ imaging traits, including brain structural, diffusion, and functional MRI^1–8^, as well as cardiovascular magnetic resonance imaging (CMR)^9–11^ and abdominal MRI^12–14^. However, a limitation of common variant signals identified by GWAS is that they often reside in non-coding regions and exhibit small effect sizes, which complicates the direct derivation of biological insights or the identification of causal genes^15–19^. By focusing on rare variants in protein-coding regions of the genome, whole exome sequencing (WES) studies aim to directly identify genes of interest.

Although existing exome-wide association studies^16,17,20,21^ (ExWAS) have identified associations for some imaging traits, our knowledge of the rare variant genetic architectures of human organs and their roles in various diseases remains substantially limited. Specifically, most previous ExWAS have focused on a single organ and/or a small set of imaging traits, lacking a multi-organ perspective that simultaneously explores the genetic effects of the human brain and body. For example, Park et al.^20^ analyzed CT imaging-derived hepatic fat, Haas et al.^14^ studied liver fat based on abdominal MRI using machine learning, Jurgen et al.^21^ studied several CMR traits, and Backman, et al. ^16^ and Karczewski et al.^17^ included brain MRI traits in their UK Biobank (UKB) WES studies of a wide range of phenotypes. Therefore, analyzing more and refined imaging traits, spanning multiple organs, in larger sample sizes will yield deeper insights into the genetic effects of rare variants across the whole body and shared genetic regulation across organs^10^.

Here we conducted ExWAS for 596 MRI traits derived from the brain, heart, liver, kidney, and lung (**Table S1**) of over 50,000 participants from the UKB study. We used an internal discovery-replication design to make the best use of available data resources to identify novel rare variants and genes associated with human organ structure and function (**Fig. 1**). We evaluated various functional annotation approaches for missense variants in gene-level set-based association testing, including conventional methods such as SIFT^22^, PolyPhen2 HDIV^23^, PolyPhen2 HVAR^23^, LRT^24^, and MutationTaster^25^, as well as the deep learning-based method AlphaMissense^26^. We compared our results with previous GWAS on the same set of MRI traits in order to provide shared common and rare variant evidence for a gene’s involvement in a trait. We also used burden heritability regression (BHR)^27^ tests to characterize the genetic architecture for ultra-rare coding variants (minor allele frequency [MAF] < 1 × 10^-4^) across functional classes. In summary, our study identified novel exome-wide associations for multi-organ structure and function, providing a valuable source of evidence that could be useful in drug discovery and clinical therapeutics. These findings may also enhance our understanding of the complex interrelations between the human organs, health, and disease.

**Fig. 1.**
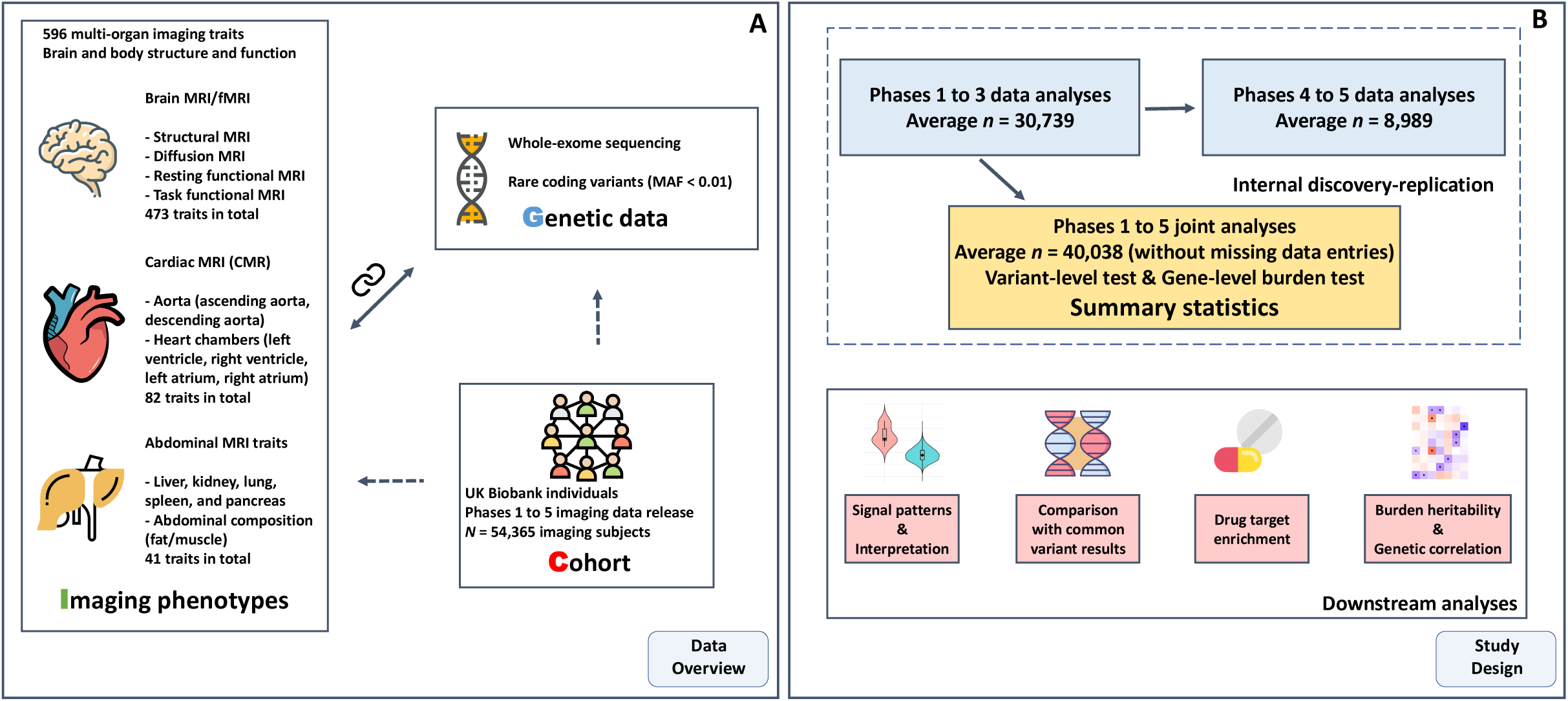
Data overview and study design. **(A)** An overview of the data included in our study. For imaging phenotypes, we made use of a broad range of multi-organ imaging phenotype data including brain imaging traits such as regional brain volume from structural MRI, diffusion tensor imaging (DTI) parameters from diffusion MRI, and functional connectivity and activity traits from resting and task fMRI; CMR traits such as measurements of the aorta (ascending/descending aorta) and heart chambers (left ventricle, right ventricle, left atrium, and right atrium); abdominal MRI traits such as abdominal organ traits (liver, kidney, lung, spleen, and pancreas) and abdominal composition measurements of fat and muscle. For genetic data, we included whole exome sequencing data and focused on rare coding variants (minor allele frequency [MAF] < 0.01). Only European individuals with imaging traits were included in the present study (*n* = 54,365). **(B)** An overview of our study design. We adopted an internal discovery-replication procedure and finalized the summary statistics based on the joint sample (phases 1 to 5 sample). Specifically, we conducted association tests on phases 1 to 3 sample, phases 4 to 5 sample, and phases 1 to 5 sample respectively. We examined (i) whether the detected signals from phases 1 to 3 sample had concordant directions and remained significant in phases 4 to 5 sample; (ii) whether the detected signals from phases 1 to 3 sample had concordant directions and obtained stronger evidence (smaller *P-*values) in the joint sample. Based on this validated procedure, the significant results and other downstream analyses used the summary statistics generated from this joint sample.

## RESULTS

### Overview of variant-level associations with multi-organ MRI traits

Based on European individuals from UKB phases 1 to 5 MRI data (released up to late 2023, average *n* = 40,038; **Fig. 1**), we identified 107 rare (MAF < 0.01) variant-level associations (**Table S2**) using a conservative *P*-value threshold of 2.8 × 10^-10^ (Bonferroni adjusted for all variant-trait tests with minor allele count [MAC] > 5 as 0.05/178,280,016, Methods) (**Fig. 2A**). Only predicted loss-of-function variants (pLoF) and missense variants were included in our study. There were 75 gene-trait pairs between 24 unique genes and 62 MRI traits, including 3 abdominal MRI traits, 7 CMR traits of the heart and the aorta, 10 brain structural MRI traits (regional brain volumes), as well as 42 brain diffusion MRI traits (diffusion tensor imaging [DTI] parameters) (**Fig. 2B**). Additionally, 138 associations (92 unique genes and 107 MRI traits) showed suggestive evidence, surviving a more liberal *P*-value threshold of 1 × 10^-8^ (**Table S3**, Methods). These associations spanned across all categories of MRI traits (**Table S2**). As expected, larger effect sizes were linked with lower MAF, which revealed the process of negative selection^17,28,29^ (**Fig. 2C**).

**Fig. 2.**
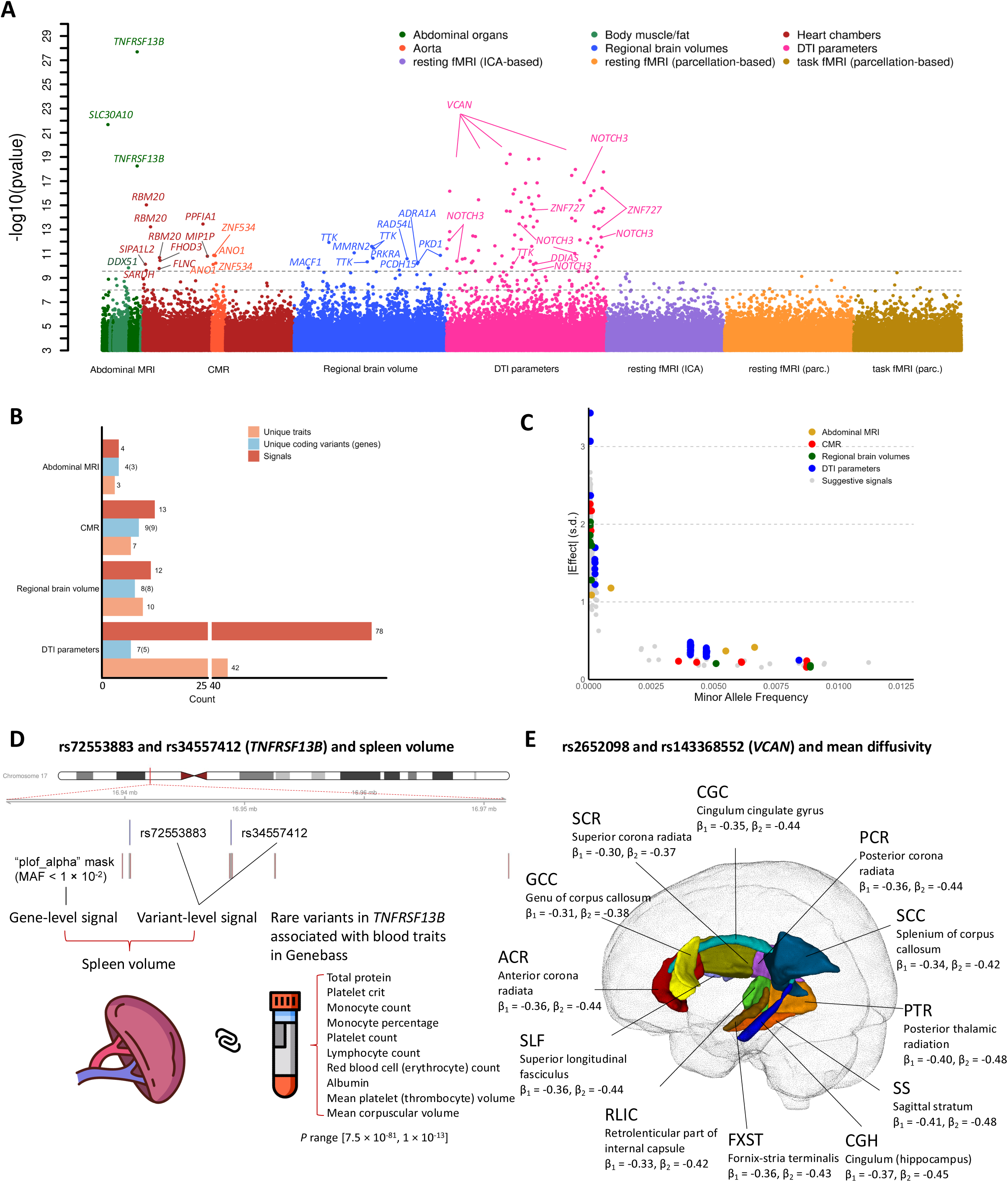
Results of variant-level association tests. **(A)** Manhattan plot for variant-level association tests across all 596 imaging phenotypes. Only coding variants of interest were included in the variant-level association tests and associations with *P* < 1 × 10^-3^ were plotted. The dashed lines indicate the variant-level significant *P*-value threshold at 2.8 × 10^-10^ as well as the suggestive *P*-value threshold at 1 × 10^-8^. The *x*-axis shows all the categories of our imaging phenotypes, while we further separated the abdominal MRI traits into abdominal organs and body/fat composition, and separated CMR traits into heart chambers and aorta areas based on the specific traits. Significant coding-variant-level associations (*P* < 2.8 × 10^-10^) were labeled to the corresponding genes. For DTI parameters, the unlabeled signals all belong to *VCAN* gene. **(B)** Summary count for the number of signals, unique variants (unique genes in the parentheses), and unique traits within each imaging category that has significant variant-level associations. **(C)** The scatter plot for all the variant-level significant results (colored by imaging categories) and suggestive results (in grey color). The *x*-axis shows the minor allele frequency of each association and the *y*-axis is the absolute value of the corresponding genetic effect size in s.d. units. **(D)** Graphical illustration of associations between two missense variants (rs72553883 and rs34557412) in *TNFRSF13B* and spleen volume. The rug plot at the first row demonstrates the locations of these two missense variants. The rug plot at the second row indicates the locations of rare variants in the ‘plof_alpha’ gene burden model, since gene-based burden test also revealed the association between *TNFRSF13B* and spleen volume. The summary statistics (*P-*values) of exome-wide associations between *TNFRSF13B* and blood traits based on burden tests were downloaded from Genebass^17^. **(E)** Visualization of the associations between two missense variants (rs2652098 and rs143368552) in *VCAN* and 12 mean diffusivity traits (β_1_ is the effect size in s.d. of rs2652098 and β_2_ is the effect size in s.d. of rs143368552).

Due to the lack of independent data sources and the inherent rarity of the variants, identifying rare variant associations typically requires large sample sizes, and findings are difficult to fully replicate. To assess the reliability of our findings and maximize the use of available data, we adopted a procedure that includes internal replication and joint analysis^30^ (**Fig. 1** and Methods). First, we limited our discovery dataset to include only individuals from phases 1 to 3 of MRI release (average *n* = 30,739). This subset yielded 59 associations that passed the Bonferroni threshold of *P* < 3.5×10^-10^ (Bonferroni adjusted for all variant-level tests with MAC > 5 in this subset, 0.05/142,935,657, Methods) (**Table S4**). Among these 59 associations, 56 had mutations in the independent sample from phases 4 and 5 of the MRI release. Of these, 47 (84% = 47/56) passed the *P*-value threshold of 2.9 × 10^-2^ (Benjamini-Hochberg false discovery rate [FDR] at the 0.05 level), and all had consistent effect size directions as those in the phases 1 to 3 data. Furthermore, 84.7% (= 50/59) of these associations had smaller *P*-values in the combined phases 1 to 5 sample, and all of them had consistent effect directions. Variants with decreased *P*-values in the joint analysis implied similar effects of the two sub-cohorts, and we found that the 50 signals that had smaller *P*-values in the combined sample indeed included all the 47 signals that were replicated in the independent phases 4 and 5 sub-cohort (**Table S4**). Overall, this internal replication analysis showed the robustness and validity of our data results. At least 84% of the 107 variant-level associations identified in the UKB phases 1 to 5 sample could potentially be replicated at the 5% FDR level, should a replication dataset become available. In the following two sections, we highlighted some interesting findings across different organs. The complete list of all these 107 variant-level associations is presented in **Table S2**.

### Variant-level tests identified novel associations for hepatic and spleen MRI traits

To our knowledge, associations between rare variants and abdominal MRI-derived traits were only studied in Hass et al.^14^, where the focus was restricted to liver fat and only 18,103 subjects were included. Another study^20^, with a similar focus on hepatic fat trait, used CT imaging data from 9,594 individuals to study the exome-wide associations. Thus, the roles of rare variants in a wider range of abdominal MRI traits were largely unexplored. In our variant-level analysis, we identified associations between spleen volume and two missense variants in *TNFRSF13B* (rs72553883, effect_org_ = 0.025 L, effect = 0.37 s.d. units, 95% CI = [0.29, 0.45], *P* = 5.6 × 10^-19^, and rs34557412, effect_org_ = 0.028 L, effect = 0.42 s.d. units, 95% CI = [0.34, 0.49], *P* = 2.0 × 10^-28^), while the latter one was the top hit among all variant-level associations. *TNFRSF13B* is a well-known risk gene of common variable immunodeficiency and mutations in *TNFRSF13B* typically occurred in patients who developed splenomegaly^31–33^. In addition, the missense variant rs72553883 was associated with a series of blood-related traits including platelet, myeloid white cell, and lymphoid white cell indices in a previous GWAS^34^ using genotyping array data while similar associations between rare variants in *TNFRSF13B* and multiple blood biomarkers were also observed in Karczewski et al.^17^ using exome data (**Fig. 2D**). Consistent with these results on spleen abnormalities derived from immunodeficiency and the central role of the spleen in blood filter and blood cell turnover, our finding points out a direct association between spleen volume and missense mutations in *TNFRSF13B*. Another missense variant in the manganese transporter *SLC30A10* (rs188273166, effect_org_ = 63.72 milliseconds^35,36^ [ms], effect = 1.18 s.d. units, 95% CI = [0.94, 1.42], *P* = 2.1 × 10^-22^) was associated with higher corrected T1 liver iron. *SLC30A10* is known to be involved in maintaining the manganese level^37^. This association aligned with recent GWAS that reported the role of rs188273166 in hypermagnesemia symptoms and a *SLC30A10*-targeted study found its positive effect on corrected T1 liver iron^38,39^. Interestingly, we also identified a missense variant in *DDX51* associated with the fat-free muscle volume of right posterior thigh (rs200735214, effect_org_ = −0.87 L, effect = −1.09 s.d. units, 95% CI = [−1.42, −0.76], *P* = 1.5 × 10^-10^), which constitutes the only signal that passed our stringent *P* value threshold for human muscle/fat composition traits. Notably, at a more permissive *P*-value threshold at 1 × 10^-8^ as our suggestive evidence, we observed that rs200735214 in *DDX51* was also associated with the fat-free muscle volume for total thigh (effect_org_ = −2.26 L, effect = −0.90 s.d. units, 95% CI = [−1.19, −0.60], *P* = 1.95 × 10^-9^) among other signals for human muscle/fat composition traits such as *PLIN4* and posterior thigh muscle fat infiltration (effect_org_ = −0.95%, effect = −0.40 s.d. units, 95% CI = [−0.53, −0.27], *P* = 1.2 × 10^-9^ for the left; effect_org_ = −0.91%, effect = −0.38 s.d. units, 95% CI = [−0.51, −0.26], *P* = 4.2 × 10^-9^ for the right). *DDX51*, a member of the DEAD-box helicase family, plays a crucial role in ribosomal RNA processing and is essential for ribosome biogenesi^40,41^. Although no direct link between *DDX51* and muscle function has been reported previously, the strong association identified in our exome data highlights its potential involvement in muscle composition traits. Meanwhile, *PLIN4* encodes a protein known as Perilipin 4, which belongs to the perilipin family of proteins that coat lipid droplets and involve in regulating lipid metabolism^42^. Perilipins are essential for the proper storage and release of lipids within cells. In particular, *PLIN4* has been shown to be highly expressed in skeletal muscle and is found at periphery of skeletal muscle fibers^42^, consistent with our findings.

### Variant-level tests identified novel associations for brain and heart MRI traits

We highlight some rare variants associations with regional brain volumes, brain DTI parameters, and CMR traits of the heart and aorta. A missense variant in *ADRA1A* was associated with the volume of the left ventral diencephalon (rs771722367, effect_org_ = 683.56 mm^3^, effect = 1.28 s.d. units, 95% CI = [0.90, 1.67], *P* = 6.5 × 10^-11^), though the *P* value for this variant and right ventral diencephalon was *P* = 3.6 × 10^-8^, which did not pass our liberal threshold at 1 × 10^-8^. *ADRA1A* was consistently reported to be associated with schizophrenia^43,44^ while enlarged brain regional volume of the ventral diencephalon area in patients with schizophrenia was previously observed^45^. The ventral diencephalon area was also associated with a missense variant in *PKD1* (rs1181041827, effect_org_ = 923.88 mm^3^, effect = 1.73 s.d. units, 95% CI = [1.21, 2.24], *P* = 5.2 × 10^-11^ for right ventral diencephalon; effect_org_ = 945.24 mm^3^, effect = 1.77 s.d. units, 95% CI = [1.26, 2.29], *P* = 1.4 × 10^-11^ for left ventral diencephalon). For DTI parameters, two missense variants (rs2652098 and rs143368552) in Versican (*VCAN*) contributed the largest number of associations (63 in total, effect range = [−0.48, 0.42] s.d. units, *P* < 2.8 × 10^-10^). **Figure 2E** visualizes the associations between these two missense variants and 12 mean diffusivity traits. Common variants in *VCAN* have been extensively associated with white matter traits in previous GWAS^2,5,8,46–48^. *VCAN* plays a pivotal role in various neural processes, which may influence the pathophysiology of neurological disorders such as multiple sclerosis^49–51^. Another missense variant rs201680145 in *NOTCH3* was also associated with multiple DTI parameters (effect range = [−1.50, 1.70] s.d. units, *P* < 2.5 × 10^-10^). Previous studies have revealed the role of *NOTCH3* in white matter hyperintensities and several neurodegenerative diseases^52–54^.

We found associations between a missense variant rs189569984 in *RBM20* and three CMR traits, including left ventricular end-systolic volume (LVESV), right ventricular end-systolic volume (RVESV) (effect_org_ = −3.77 mL, effect = −0.20 s.d. units, 95% CI = [−0.25, −0.15], *P* = 9.1 × 10^-16^ for LVESV; effect_org_ = −3.37 mL, effect = −0.16 s.d. units, 95% CI = [−0.21, −0.11], *P* = 2.1 × 10^-11^ for RVESV), and left ventricular ejection fraction (LVEF) (effect_org_ = 1.49 %, effect = 0.24 s.d. units, 95% CI = [0.18, 0.30], *P* = 6.0 × 10^-14^). Mutations in *RBM20* were previously known to be related to cardiovascular diseases including heart failure and dilated cardiomyopathy^55–57^, and were associated with LVEF in a previous GWAS on CMR traits^9^. Our rare variant analysis prioritized *RBM20* and provided additional evidence for its role in regulating heart structure and function. Furthermore, we found that ascending aorta maximum/minimum areas were associated with a missense variant in *ANO1* (rs201870990, effect_org_ = 41.95 mm^2^, effect = 0.22 s.d. units, 95% CI = [0.15, 0.29], *P* = 7.6 × 10^-11^for maximum; effect_org_ = 42.05 mm^2^, effect = 0.23 s.d. units, 95% CI = [0.16, 0.29], *P* = 1.4 × 10^-11^ for minimum). *ANO1* was among the loci identified by a recent GWAS^58^ on ascending aorta diameter (see Figure 1 in their study^58^) but was not pointed out or discussed explicitly. Indeed, *ANO1* was also reported to be effective in preventing cardiac fibrosis and may be a potential target for therapy^59,60^. However, future research might elucidate its role in aortic development and/or geometric remodeling. In summary, our analysis of rare variants directly prioritized and implied a small set of genes related to human organs. Due to the rarity of these variants and their lack of linkage disequilibrium (LD) with common variants, they could not be effectively studied in previous GWAS that focused on common variants.

### Gene-based burden tests identified complementary signals

Gene-based burden tests enable us to study the collective effects of rare variants within a gene, thus boosting the power. However, the involved burden models pose challenges to the adjustment for multiple testing since they have unknown correlated structure. Thus, we prioritized a *P*-value threshold at 1 × 10^-9^ based on two empirical null distribution^61,62^ (Methods). Using the same dataset as in variant-level tests, we identified 224 significant associations in gene-based tests (*P* < 1 × 10^-9^, Methods; **Table S5** for all the significant results). As different burden models or MAF cutoffs may implicate the same gene-trait associations (Methods), we further summarize the nonredundant results of 67 unique gene-trait pairs^61^ (involving 26 genes and 57 MRI traits) in **Table 1** (a more comprehensive version is provided in **Table S6**). For suggestive evidence, we additionally put all the associations that passed a more relaxed *P*-value threshold at 1 × 10^-8^ in **Table S7**. Manhattan plot for all the gene-level associations were presented in **Figure 3A**, across all MRI phenotype categories. Consistent with observations in previous ExWAS^17,62^,we identified 21 additional genes, highlighting the power of gene-based burden tests in rare variant association studies. In particular, we found 3 genes and 18 gene-trait associations for brain functional MRI (fMRI) traits, which did not have any signals in variant-level tests. To evaluate the reportability of our findings, we wanted to follow a similar procedure in variant-level tests to perform an internal replication. However, many rare mutations, particularly those with a MAF of less than 0.01%, accounted for a large proportion of our discoveries but were not observed in the smaller independent dataset from phases 4 to 5 (average *n* = 8,989). Therefore, we only examined whether the associations in the combined phases 1 to 5 sample had smaller *P*-values than those in the phases 1 to 3 data (Methods). When we restricted the discovery sample to individuals in phases 1 to 3, there were 96 significant associations for 25 unique gene-trait pairs (*P* < 1 × 10^-9^, **Table S8**). Within these 96 associations, 79.2% (76/96, 19 gene-trait pairs) had smaller *P*-values in the combined phases 1 to 5 sample, all of which had concordant directions of effects (**Table S8**). We highlighted these 19 gene-trait pairs in **Table 1** and discussed some of the 67 gene-trait pairs in the following two sections.

**Fig. 3.**
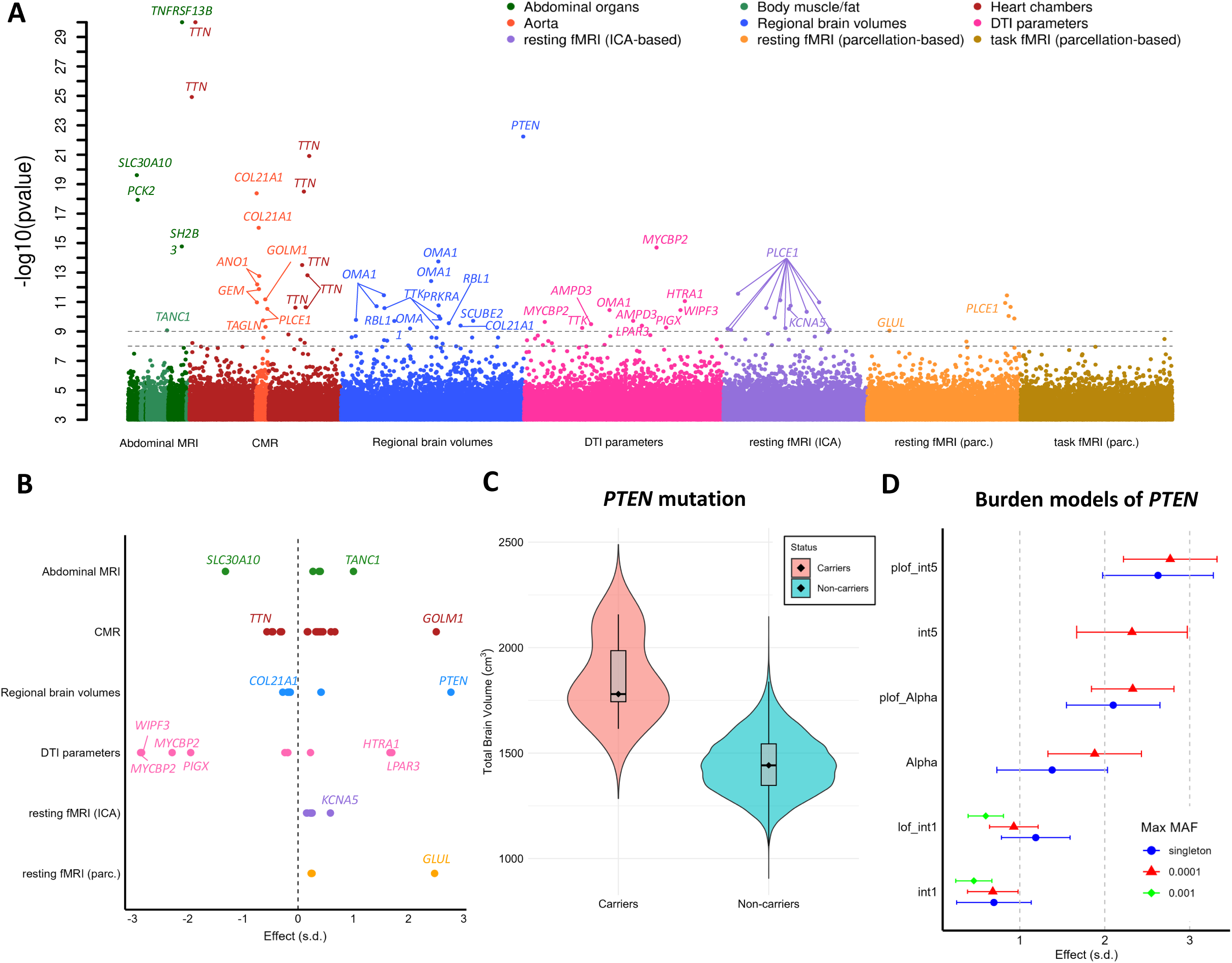
Results of gene-level burden tests. **(A)** Manhattan plot for gene-level burden tests across all 596 imaging phenotypes. For significant gene-trait pairs appearing in multiple burden models, we plotted the association with the smallest *P*-value. The dashed lines indicate the gene-level significant *P*-value threshold at 1 × 10^-9^ as well as the suggestive *P*-value threshold at 1 × 10^-8^. The *y*-axis is capped at 1 × 10^-30^ and only gene-trait pairs with *P* < 1 × 10^-3^ were included. The color legend and *x*-axis are same as the Manhattan plot for variant-level associations in Figure 2A. Significant genes were labeled. **(B)** Effect sizes for all non-redundant significant gene-trait pairs (*n* = 67, corresponding to **Table 1**) within each imaging category. The dashed line indicates an effect size of zero. We labelled all the genes that had the largest positive and negative effects within each imaging category and genes that had an absolute effect size greater than 1.5. **(C)** Distribution of total brain volume (in cm^3^) of mutation carriers versus non-carriers of *PTEN*. To illustrate, we select the burden model with the smallest *P*-value, that is “plof_int5”, which means the aggregation of pLoF variants and missense variants with a minor allele frequency (MAF) cutoff at 1 × 10^-4^. **(D)** The effect size estimates and the corresponding 95% confidence intervals across different burden models of *PTEN*. The color indicates distinct MAF cutoffs. The *y*-axis includes all the burden models (Methods). Only burden models with minor allele count > 5 (after aggregation) were tested (Methods) and plotted.

### Genes associated with abdominal, brain, and heart MRI traits in burden tests

As shown in **Table 1**, we identified 5 genes for abdominal MRI traits, 7 for CMR traits, 7 for regional brain volumes, 8 for brain DTI parameters, and 3 for brain fMRI traits. **Figure 3B** highlights the genes with large effect sizes. Below we highlight several interesting findings for each trait category.

We found that abdominal MRI signals were sparse and robust, with all 5 signals from the combined phases 1 to 5 sample also present in the phases 1 to 3 data. These signals were the top-ranking signals among all imaging categories. The highest hit among all the gene-level signals was the association between spleen volume and *TNFRSF13B* when aggregating the effects of pLoF and Alpha damaging (Methods) missense variants (effect_org_ = 0.027 L, effect = 0.38 s.d. units, 95% CI = [0.29, 0.48], *P* = 1.7 × 10^-55^). In addition to *TNFRSF13B*, we observed that *SH2B3* was also associated with spleen volume (effect_org_ = 0.025 L, effect = 0.40 s.d. units, 95% CI = [0.35, 0.45], *P* = 1.7 × 10^-15^), where spleen abnormalities (e.g. splenomegaly) and associated blood traits have been previously reported^63–66^. For muscle measurements, we found a novel association between fat-free muscle volume of the posterior thigh and *TANC1* (effect_org_ = −1.06 L, effect = −1.32 s.d. units, 95% CI = [−1.74, −0.90], *P* = 8.5 × 10^-10^). *TANC1* had genetic overlaps with the identified loci in previous GWAS on heel bone mineral density^67^ and its role in muscle development and rhabdomyosarcoma has been discussed in previous studies^68,69^.

As shown in **Figure 4A**, *TTN* was associated with 8 CMR traits, including LVESV (effect_org_ = 7.34 mL, effect = 0.40 s.d. units, 95% CI = [0.32, 0.47], *P* = 1.2 × 10^-25^), LVEF (effect_org_ = −3.48 %, effect = −0.56 s.d. units, 95% CI = [−0.66, −0.47], *P* = 2.1 × 10^-32^), as well as global and regional peak circumferential strain measurements (effect range = [0.32, 0.60] s.d. units, *P* range = [1.2 × 10^-21^, 2.4 × 10^-11^]). These results make sense as *TTN* is a well-known gene associated with cardiac structure^9^ and cardiovascular diseases such as heart failure^70^, dilated cardiomyopathy^71,72^, atrial fibrillation^73^, supraventricular tachycardia, and mitral valve disease^21^. In addition to the previously known exome-wide association between *TTN* and LVESV^21^, our findings suggest a broader influence of this gene on cardiac structure and function, using a larger sample size and an expanded set of CMR traits.

**Fig. 4.**
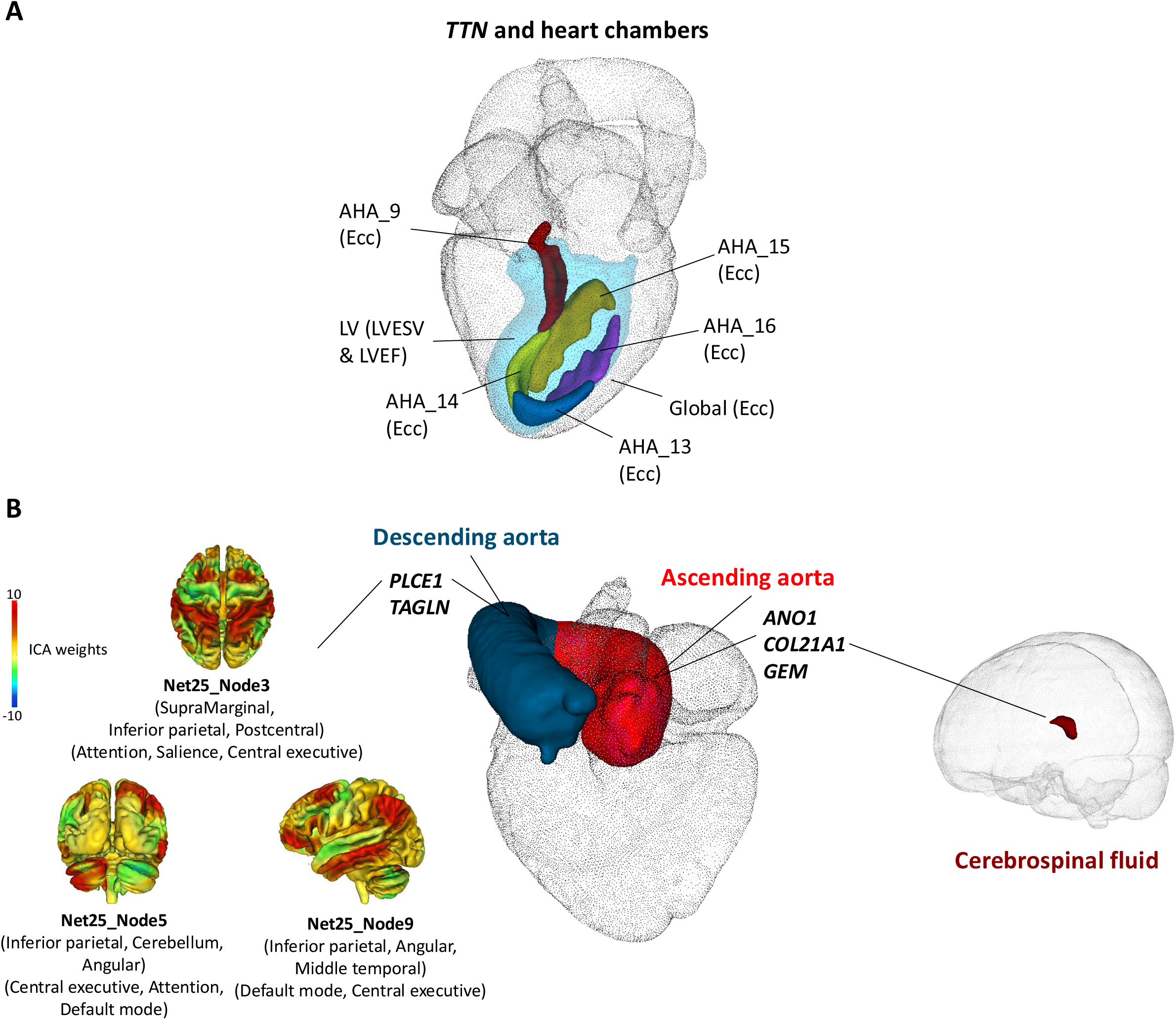
Heart and aorta-related associations and heart-brain connections. **(A)** Graphical illustration of 8 CMR traits of left ventricle (LV) associated with *TTN*. LVESV, left ventricular end-systolic volume; LVEF, left ventricular ejection fraction; and Ecc, peak circumferential strain, including both global (“Global”) and regional traits (such as “AHA_9”). **(B)** Five genes (*ANO1, COL21A1, GEM, PLCE1,* and *TAGLN*) associated with ascending aorta and descending aorta areas. *PLCE1* was also associated with brain fMRI traits. Three ICA-based functional activity traits (Net25_Node3, Net25_Node5, and Net25_Node9) associated with *PLCE1* are illustrated, with their major brain regions and networks labeled. The color represents the weight profile of the ICA node. In addition, *COL21A1* was associated with cerebrospinal fluid volume.

We would like to highlight our exome-wide associations with CMR traits of aorta. Notably, three genes (*ANO1, COL21A1, GEM*) were associated with both maximum and minimum area of ascending aorta, while *PLCE1* was associated with maximum and minimum area of descending aorta (**Fig. 4B**). In particular, both *COL21A1* and *GEM* lower the size of the ascending aorta (effect_org_ range = [−61.16, −54.95] mm^2^, *P* range = [1.7 × 10^-13^, 6.4 × 10^-13^] for *COL21A1* and effect_org_ range = [−87.68, −87.06] mm^2^*, P* range = [1.3 × 10^-12^, 1.06 × 10^-11^] for *GEM,* respectively). Rare variants in *COL21A1* and *GEM* were reported to be associated with higher pulse blood pressure in a previous study for blood pressure^74^. A smaller aorta leads to a wider pulse pressure because it increases aortic characteristic impedance, which is highly dependent on aortic diameter. Our results on these aorta area-lowering genes further provide the evidence for the underlying mechanisms of previous observations. Moreover, *GEM* has a role in regulating cardiac activities^75^ and is related to heart failure^75,76^. Another study^77^ pointed out that such effects of *GEM* could potentially be used as a gene therapy target in treating ventricular arrhythmias and heart failure. Our novel discovery indicated that *GEM*’s regulatory effects may also impact the structural integrity of the ascending aorta, thus improving our understanding of the role of *GEM* in cardiovascular health. In contrast, missense variants in *ANO1* have positive effect. The role of *ANO1* has been discussed in our variant-level analysis and we recaptured these associations again by aggregating the missense variants in *ANO1* using burden test (effect_org_ range = [32.43, 32.79] mm^2^, *P* range = [1.7 × 10^-13^, 6.4 × 10^-13^]), suggesting the similar effects and aligned directions across a group of missense variants within *ANO1*. In addition, *TAGLN* was also associated with descending aorta minimum area (effect_org_ = 62.28 mm^2^, effect = 0.67 s.d. units, 95% CI = [0.46, 0.88], *P* = 4.91 × 10^-11^) while its association with descending aorta maximum area was observed at a more liberal *P* value threshold at 1 × 10^-8^ (**Table S7**). *GOLM1* was found to be associated with descending aorta distensibility (effect_org_ = 2.85mmHg, effect = 2.5 s.d. units, 95% CI = [1.77, 3.24], *P* = 2.93 × 10^-11^). A recent study demonstrated that *GOLM1* may present a potential therapeutic target for treat sepsis-induced cardiac dysfunction in animal models^78^. *GOLM1* was also related to Alzheimer’s disease and the implied cognitive deficits^79^. However, its role in aortic wall remodeling requires further study. Using GTEx data resources^80^, we found that four (*ANO1, COL21A1, GEM, TAGLN*) out of five genes (*ANO1, COL21A1, GEM, TAGLN, PLCE1*) associated with ascending/descending aorta areas had moderate to high expression and overexpressed in aorta while *PLCE1* had high expression in artery tissues in general (Methods), which further supports their roles in regulating the structure of aorta.

For brain MRI traits, we highlight in **Figures 3C** and **3D** the strongest association between *PTEN* and total brain volume with extraordinary effect size when aggregating pLoF and missense variants (effect_org_ = 386703.40 mm^3^, effect = 2.77 s.d. units, 95% CI = [2.22, 3.32], *P* = 5.8 × 10^-23^). The distribution of total brain volume in cm^3^ for *PTEN* mutation carriers versus non-carriers is provided in **Figure 3C**. Notably, as illustrated in **Figure 3D**, different burden masks would yield associations with various strengths, and we observed that the signal became stronger if we aggregate ultrarare (MAF < 1 × 10^-4^) pLoF variants and damaging missense variants in *PTEN*, which may not be observed and testable in the previous ExWAS^16,17^ with a sample size smaller than ours. In a recent ExWAS^81^, *PTEN* was associated with autism spectrum disorder. Indeed, *PTEN* is a well-known gene that has been consistently linked to autism spectrum disorder^82–85^, brain development^86–88^, and brain clinical phenotypes including brain overgrowth^89–91^, with an enlarged brain volume representing an associated phenotype in these conditions. To our knowledge, no previous GWAS or ExWAS has linked *PTEN* to any brain MRI measurements, thus our analysis provided direct evidence connecting rare variants in *PTEN* with total brain volume. Another gene *OMA1* was observed to be associated with 7 regional brain volume traits including superior frontal gyrus, lateral orbitofrontal cortex, and precentral gyrus for both sides of the brain and right superior parietal lobule (effect_org_ range = [−524.83, −171.02] mm^3^ units, *P* range = [1.77 × 10^-14^, 6.35 × 10^-10^]). Notably, *PTEN* and *OMA1* regulate the *PTEN*-induced kinase 1 (*PINK1*) and thus may be useful in preventing epileptogenesis^92^ while a protective role of *OMA1* in neurodegeneration^93–95^ was previously reported. The remarkably strong association *PTEN* demonstrate that even within a general healthy cohort (as opposed to disease-focused or case-control studies), we can identify biological links between genes associated with neurodevelopmental disorders using MRI data.

For DTI parameters, in contrast to variant-level analysis where missense variants in *VCAN* accounted for the most associations, our gene-based burden tests revealed a more diverse range of effects on white matter, with 8 distinct genes accounting for 10 gene-trait associations. For brain fMRI traits, *PLCE1*, *GLUL*, and *KCNA5* were identified to be associated with 18 phenotypes, where *PLCE1* contributed to 16 of them. We found that many gene-trait pairs would not have been identified if singleton burden models or the recent deep learning-based AlphaMissense^26^ burden models were not included. We will provide more detailed discussions about these interesting associations in the following sections.

Overall, gene-based burden tests examined the aggregated effects of rare variations within genes and advanced our understanding of the underlying genetic dispositions across the abdomen, heart, and brain organs. Our novel findings prioritized a set of rare genes previously unknown to be associated with human organs. We further discussed examples of genetic overlaps between these MRI signals and other health-related phenotypes, which may help understand complex diseases by providing exome-wide insights into the genetic mechanisms underlying multiple human organs including abdominal, cardiac, and neurological pathologies.

### Pleiotropic effects of *PLCE1* and *COL21A1* on brain and heart MRI traits

In our gene-based burden tests, *PLCE1* was associated with the largest number of phenotypes including 16 brain fMRI traits (including 5 traits from parcellation-based approach^96^ and 11 traits from whole brain spatial independent component analysis [ICA]^97–99^) and 2 CMR traits (descending aorta maximum/minimum areas) (**Fig. 4B**). In addition, *COL21A1* was associated with both brain cerebrospinal fluid (CSF) volume and ascending aorta maximum/minimum areas (**Fig. 4B**). We found that a recent GWAS^100^ discovered the associations of descending and ascending aorta areas separately with *PLCE1* and *COL21A1* while *PLCE1* was also found in previous GWAS^3,5,101^ for brain fMRI. *PLCE1* encodes a phospholipase (PLCε) that catalyzes the hydrolysis of phosphatidylinositol-4,5-bisphosphate to generate inositol 1,4,5-triphosphate and diacylglycerol. They are common second messengers regulating multiple cellular processes, including cell activation, growth, differentiation, and gene expression^102^. Interestingly, prior mechanistic study identified a causal role for PLCε on the development of thoracic dilation^103^ and dissection^104^. Similarly, *COL21A1* encodes the alpha-1 chain of collagen 21, a known extracellular matrix component in the arterial wall, secreted by vascular smooth muscle cells. While the previous GWAS^100^ suggested distinct genetic bases between the ascending and descending aorta and linked aortic traits to brain small vessel disease, it did not establish direct associations of these genes with other brain MRI traits. Our results provide exome-wide evidence highlighting the associations of *PLCE1* and *COL21A1* with both brain and heart MRI traits. Additionally, a previous study suggested that the observed GWAS effects of *PLCE1* on fMRI traits may be blood-derived^101^. Our exome-wide evidence supporting *PLCE1*’s role in both aortic and fMRI traits aligns with this hypothesis.

The connection between the heart and brain has increasingly garnered attention^10,105^. Moreover, the role of hemodynamic role of the aorta involved the cushioning of pressure and flow pulpability caused by the intermittent ventricular ejection, and an impaired cushioning function for changes in aortic stiffness and/or diameter has been identified as a key mechanism for target organ damage, including the brain and the heart^106^. The associations we identified, involving rare coding mutations in the same genes (*PLCE1* and *COL21A1*) linked to heart, aortic and brain structure and function, suggest that similar biological mechanisms may underlie the shared pathways of these organs that influence both cardiac and neurological health.

### Novel associations uniquely identified by singletons and AlphaMissense

Singleton variants are those observed only once in the study cohort. AlphaMissense^26^ is a recent deep learning-based method that offers a novel approach to annotating damaging missense variants. Singleton burden model captured the rarest category in our association tests and may be of special interest^16,107^, and the incorporation of AlphaMissense offered novel genetic findings in our study. Therefore, we would like to highlight the contributions of these two groups of burden models in this section. In our gene-based burden tests, 10 gene-trait pairs would not be detected without the singleton burden models or including AlphaMissense as part of our annotation resources. Among these signals, 8 of them even did not have any counterparts that passed the relaxed *P* < 1 × 10^-8^ threshold (**Table S9**).

Singleton damaging missense variants in glutamate-ammonia ligase *GLUL*, which encodes the glutamine synthetase protein, were associated with functional connectivity of the somatomotor and secondary visual networks (effect = 2.48 s.d. units, 95% CI = [1.68, 3.27], *P* = 8.9 × 10^-10^). *GLUL* knockout mice were reported to be neuronally affected in multiple regions including somatosensory and visual cortices^108^. Our singleton analysis provides consistent evidence in human genetics. In addition, a recent study suggested that visual-somatosensory integration may be a new biomarker for preclinical Alzheimer’s disease^109^. Together, rare mutations in *GLUL* may play a potential role in human neurodegenerative diseases^110–113^. Another gene, *KCNA5* related to potassium voltage-gated channel, was associated with functional activity in the subcortical-cerebellum network when we combined the rare pLoF variants and Alpha damaging missense variants (effect = 0.59 s.d. units, 95% CI = [0.41, 0.76], *P* = 2.9 × 10^-11^). Notably, voltage-gated potassium channels are essential for neurons and cardiac activities and previous GWAS have discovered the common variants mapped to *KCNA5* associated with cortical surface^7^ and thickness^114^. In addition, a previous study on the rat cerebellum model revealed strong *KCNA5* immunoreactivities in the cerebellar nuclei^115^. Consistent with these findings, we provided additional evidence for *KCNA5*’s role in brain function. Moreover, some studies also found close relationships between *KCNA5* and heart diseases including atrial fibrillation^116^ and pulmonary arterial hypertension^117^. Indeed, *KCNA5* is an approved drug target for the treatment of cardiac arrhythmias. Inspired by this additional context, we investigated whether *KCNA5* was associated with any CMR traits. Interestingly, we found that when aggregating the singleton pLoF and damaging missense variants, *KCNA5* was associated with a regional peak circumferential strain measurement (effect_org_ = 5.11%, effect = 1.08 s.d. units, 95% CI = [0.63, 1.54], *P* = 3.2 × 10^-6^, **Table S1**) at a *P*-value threshold of 1.74 × 10^-5^ (0.05/2,870, Bonferroni adjusted for 82 CMR phenotypes across all the 7 variant function classes and 5 MAF classes). These discussions further suggested that multi-organ imaging genetic studies would bring about insights to the complex interplay across the brain-heart system. In summary, our results underscore the efficacy of our gene-based burden test approach and highlight the importance of using multiple MAF cutoffs and innovative annotation tools.

### Comparison with previous ExWAS for brain MRI and CMR traits

In this section, we discuss the links between our gene-based rare variant signals and existing ExWAS results on MRI traits. To our knowledge, there were no rare variant associations reported for brain fMRI or abdominal MRI traits. The studies most similar to ours are those by Backman et al.^16^, which included regional brain volumes and DTI parameters among a broad range of phenotypes, and Jurgen et al.^21^, which focused on CMR traits, while Pirruccello et al.^118^ declared no findings of rare variant associations for aorta traits. Both studies used data from the UKB cohort but with smaller sample sizes compared to our current study. Importantly, we analyzed different brain MRI and DTI parameters, extracted from raw images using our own pipeline, compared to those investigated by Backman et al.^16^. Additionally, we incorporated a broader set of CMR traits, including strain and thickness metrics, expanding upon the traits analyzed by Jurgen et al.^21^.

For regional brain volumes and DTI parameters (corresponding to STR and dMRI traits in Backman et al.^16^), 5 genes (*AMPD3*, *HTRA1*, *MYCBP2, RBL1*, *SCUBE2*) whose associations passed our stringent *P*-value threshold (*P* < 1 × 10^-9^) were also significantly associated with their brain MRI traits. For example, associations between *AMPD3* and mean MD of splenium of corpus callosum passed the stringent *P* value threshold both in our study and in Backman et al.^16^. *SCUBE2* was significantly associated with the regional volume of the left cerebellum exterior in our analysis, while its association with the volume of the cerebellum cortex in the left hemisphere was reported in their result as suggestive evidence. *MYCBP2* was associated with the external capsule in both our analysis and Backman et al.^16^. Additionally, several associations between *PLEKHG3* and multiple dMRI traits were identified in Backman et al.^16^ and two gene-trait pairs within these associations passed our relaxed threshold (*P* < 1 × 10^-8^). For CMR traits, we replicated the only exome-wide significant association between *TTN* and LVEF in Jurgen et al.^21^, while their suggestive association between *TTN* and LVESV also passed our stringent threshold. In addition, we found 7 novel associations between *TTN* and 7 peak circumferential strain metrics. We summarized the overlapping associations in these previous ExWAS studies in **Table S10**.

### Concordant evidence with GWAS signals

Associations with rare coding variants could prioritize genes among the numerous loci identified in GWAS for polygenic complex traits by providing concordant evidence^16,21^. In this section, we leveraged the previous GWAS summary statistics^1–3,10,119^ of all the same 596 imaging traits to compare the identified signals between common variants and rare coding variants (Methods). As expected, we observed convergent evidence for all categories of MRI traits. Briefly, more than half of the 174 rare coding signals (53.4% = 93/174, that is, 64/107 variant-trait associations and 29/67 gene-trait pairs) were within the 1Mb range of the independent GWAS signals, which is consistent with the observation in a previous large-scale UKB phenotype screening^16^. Based on our results, we found that variant-level associations with regional brain volumes and CMR traits were not within the neighborhood of GWAS signals, while all categories of MRI traits had at least one rare coding association within the 1Mb of GWAS signal for signals from gene-level tests.

We zoomed into these shared signals between GWAS and ExWAS to provide more detailed insights. The association between *SH2B3* and spleen volume identified in our gene-level burden test was further supported by an independent GWAS signal rs2239194, which is an expression quantitative trait locus (eQTL) in spleen tissue^80^. For DTI parameters, two missense variants (rs2652098 and rs143368552) in *VCAN* contributed the most to the variant-level associations and all these associations were within 1Mb of the GWAS signals. However, similar to findings from a previous GWAS study^48^, we did not identify any brain tissue-related eQTLs. Associations between *AMPD3* and DTI parameters of splenium of corpus callosum were also accompanied by signals from GWAS. These common variants are eQTLs of multiple brain tissues including cerebellum and spinal cord where brain white matter also presents. For fMRI traits, all the associations between *PLCE1* and 18 MRI traits (16 brain fMRI traits and 2 CMR traits) were consistent with previous GWAS results, though there were not eQTLs in brain cortex tissues. On the other hand, we found that the only signal for brain volumes that fell into the GWAS loci was the association between *COL21A1* and CSF volume. The corresponding common variants rs3857615 and rs9475654 were eQTLs in cerebellum and cerebellum hemisphere.

In addition, we identified gene-trait associations that were not reported in our previous GWAS of the same MRI traits but appeared in other studies (e.g. *RBM20* and LVEF in Pirruccello et al.^9^). We also observed that some rare coding associations in our study were linked to GWAS loci of related traits within the same phenotype cluster. For example, though both ExWAS and GWAS revealed the associations between *TTN* and multiple regional peak circumferential strain measurements, the phenotype for the signals did not exactly match. In summary, rare variant associations with MRI traits not only identified novel signals but also prioritized genes among thousands of GWAS loci. These associations provide new insights and context for understanding the genetic basis of structure and function in the human brain and body. We provided these exome-wide signals that showed convergent evidence with previous GWAS signals in **Tables S11-S12**.

### Multi-organ imaging genetics and drug targets

We used the Therapeutic Target Database (TTD)^120^ to query potential drug target genes (Methods). Among the 26 unique genes identified by gene-based burden tests, 6 genes were approved drug targets (*KCNA5* and *ANO1*) or in clinical trials (*TTK*, *GEM*, *LPAR3*, and *TNFRSF13B*). The identified genes in gene-based burden tests were significantly enriched for potential drug targets (6 out of 26, compared with 1,735 out of 17,448 genes, OR = 2.7, *P* = 0.0391, **Table S13**). Moreover, when including a broader set of potential drug genes that were documented in TTD to be reported in the literature, the enrichment became stronger (13 out of 26, compared with 3,468 out of 17,448 genes, OR = 4.0, *P* = 5.737 × 10^-4^, **Table S13**). Consistent with the procedure in a previous study^62^ and observations in other ExWAS^16,62,121^, our results suggest that rare coding variants identified in imaging genetics may facilitate the discovery of promising drug target genes^122,123^. Given the complicated nature of human organ interplay and the corresponding biological pathways involved, we would emphasize that such enrichment should be interpreted carefully as they might not directly point to the corresponding diseases and related traits for existing drugs. However, these associations with human organs could provide novel drug targets if they are supported by further evidence, and in turn these MRI traits themselves might be useful to validate the new drugs if the underlying mechanisms are well established^124^. Moreover, As endophenotypes for complex diseases, MRI traits can offer additional information on known drug targets, such as potential off-targets and/or side effects^122,125^, and also aid in drug repurposing (e.g. *ANO1* and cardiac disease as discussed in the previous section^59,60^).

### Burden heritability and genetic correlation

To investigate the rare coding genetic architecture for multi-organ MRI traits, we applied burden heritability regression (BHR)^27^ to estimate the heritability for pLoF and damaging missense variants (i.e. “int1 missense variants”, Methods), while the heritability for synonymous variants is also calculated and it serves as negative control. Designed for rare variants with MAF < 1 × 10^-3^, BHR stratifies variants based on functional annotation and MAF to allow for different variant classes to have different mean effect sizes. Given our relatively small sample size (average *n* = 40,038), we restricted our analysis to the ultra-rare MAF bin (MAF < 1 × 10^-4^) which contained most of the coding variants in our analysis (**Table S14**). As expected, ultra-rare pLoF variants consistently demonstrated higher heritability than damaging missense variants across all MRI categories (**Fig. 5A**). **Tables S15-S16** show the complete list of burden heritability for ultra-rare pLoF variants and damaging missense variants. In contrast, there is no evidence that synonymous variants have significant heritability, showing that the estimates from our results for rare variants are well calibrated (**Table S17**). Furthermore, the highest average heritability of ultra-rare pLoF variants were observed in the DTI parameters category. For ultra-rare pLoF variants, the trait had the highest heritability was the caudal anterior cingulate (*h*² = 0.012, SE = 0.0035) among regional brain volumes, the mean axial diffusivity of the body of the corpus callosum (*h*² = 0.020, SE = 0.0048) among DTI parameters, functional connectivity between the second visual and auditory networks (*h*² = 0.020, SE = 0.0047) among brain fMRI traits, regional peak circumferential strain (*h*² = 0.016, SE = 0.0053) among CMR traits, and pancreas iron (*h*² = 0.019, SE = 0.0053) among abdominal MRI traits. We note that these burden *h*² estimates are relatively large, which is similar to the high common variant heritability for many imaging traits. This in general supports an endophenotype model where genes have more direct connections with imaging phenotypes than downstream health outcomes.

**Fig. 5.**
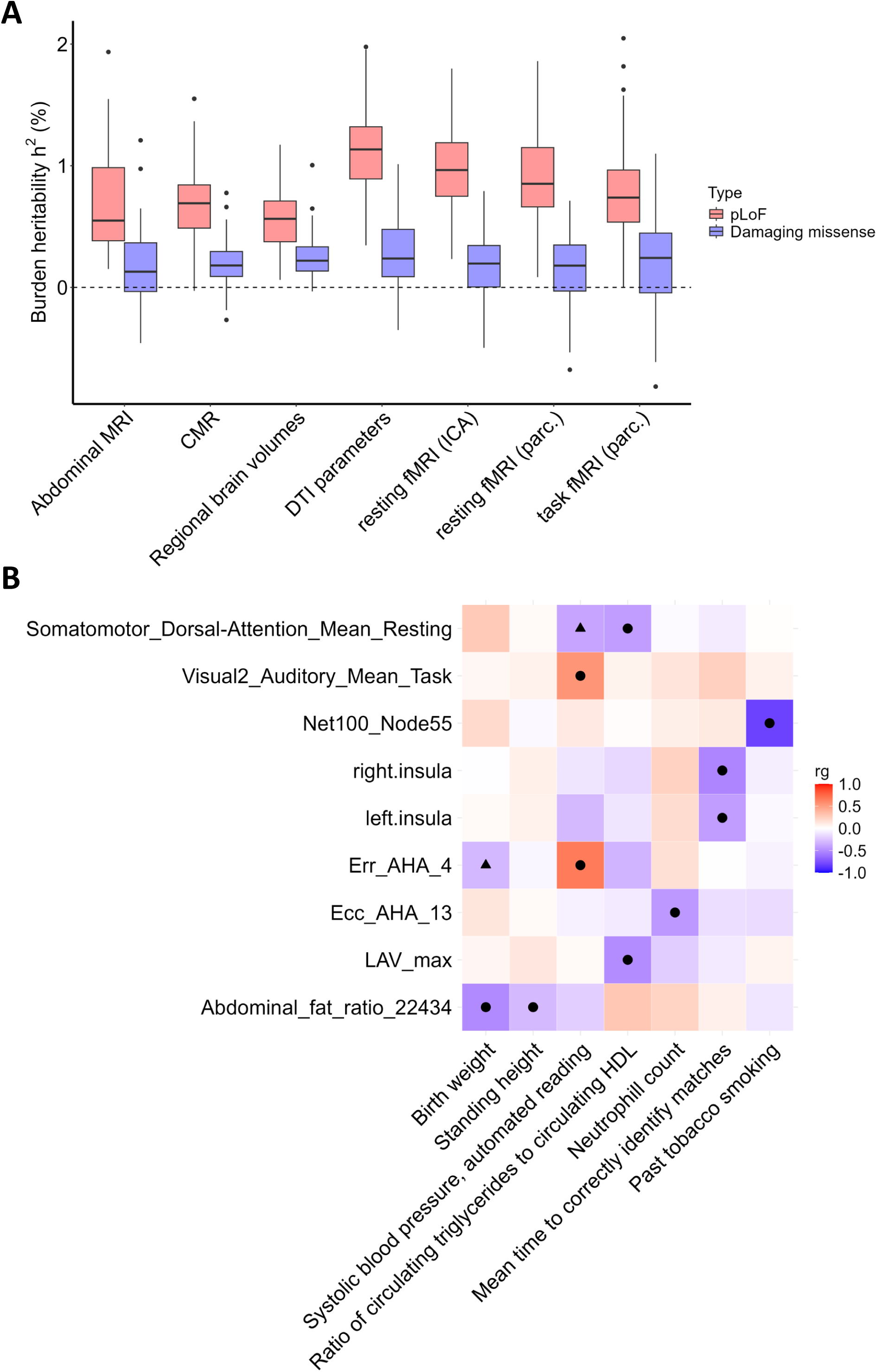
Burden heritability and genetic correlation. **(A)** The distribution of burden heritability point estimates for ultra-rare (minor allele frequency < 1 × 10^-4^) pLoF variants and damaging missense variants (i.e. the int1 damaging missense variants) across 591 multi-organ imaging traits that had a sample size larger than 10,000 (5 abdominal imaging traits were excluded due to insufficient sample size). **(B)** Burden genetic correlations of ultra-rare (minor allele frequency < 1 × 10^-4^) pLoF variants for selected imaging phenotypes and other complex traits. Only top hits are presented here. Specifically, we first selected seven trait pairs from pairs that had ten smallest *P*-values among all the trait pairs and additionally included three pairs that had smallest or second smallest *P*-values for regional brain volumes and abdominal MRI traits, which resulted in nine imaging traits across all MRI categories and seven Genebass traits. Filled circles indicate the selected top hits (*P* < 0.0032) while filled triangles indicate other pairs that passed a nominal significance threshold at *P* < 0.05. “Phenotype_ID” was used to define the selected imaging traits. Specifically, abdominal fat ratio from abdominal MRI traits, left atrium maximum volume (LAV_max), regional radial strain (Err_AHA_13) and peak circumferential strain (Err_AHA_4) traits from CMR, left and right insular volumes, functional activity trait from resting fMRI (ICA-based, Net100_Node55) traits, Visual2-Auditory network functional activity trait from task fMRI (parcellation-based), and Somatomotor-Dorsal-Attention network functional activity trait from resting fMRI (parcellation-based) traits were shown here (bottom-to-top order). **Table S1** includes more details of these phenotypes.

We further explored the burden genetic correlation pattern for ultra-rare pLoF variants between MRI traits (MAF < 1 × 10^-4^) and 186 complex traits and diseases (MAF < 1 × 10^-5^) downloaded from Genebass^17^ (**Table S18**). We focused on 67 heritable MRI traits that passed the Bonferroni adjusted threshold at *P* < 8.4 × 10^-5^ (= 0.05/596). Consequently, we estimated 12,462 (67 x 186) pairs of burden genetic correlations. We found 3 pairs that passed the MRI-multiple testing threshold at *P* < 7.5 × 10^-4^ (0.05/67) and 591 pairs passed a nominal *P*-value threshold at *P* < 0.05. Although no pairs survived the most stringent *P*-value threshold adjusted for both the imaging traits and Genebass traits at *P* < 4.0 × 10^-6^ (0.05/12,462), we observed that there were interesting pairs in the top-ranking subset for continuous traits from Genebass (**Table S19**). The results for binary traits can be found in **Table S20**. **Figure 5B** shows the selected top hits of burden genetic correlation pairs, including insular volume and reaction time^126^, regional radial strain and systolic blood pressure, left atrium maximum volume and triglycerides to high-density lipoprotein cholesterol ratio, and abdominal fat ratio and birth weight. These burden genetic correlations between MRI traits and health-related traits may reveal a shared genetic basis in ultra-rare pLoF variants. While the current MRI sample size was slightly underpowered, future larger samples may enable a more robust quantification and confirmation of these genetic links.

## DISCUSSION

We conducted a large-scale ExWAS for 596 multi-organ imaging traits including brain, heart, liver, kidney, and lung. By using both WES and MRI data from over 50,000 participants from the UKB study, we uncovered how rare coding variants contribute to human organ structure and function. The role of rare variants in regulating human organs was largely understudied. For example, in abdominal organs, a prior ExWAS focused solely on hepatic fat measurement from CT imaging^20^, while another GWAS included a broader set of traits^13^. However, with a relatively limited sample size of over 10,000, it failed to identify rare variant associations. For CMR traits, our study incorporated refined measurements of the left ventricle, right ventricle, ascending/descending aorta, and left/right atrium. These were not examined in previous ExWAS^21^ but resulted in novel discoveries in our analysis. Additionally, our results helped prioritize genes among previously identified GWAS loci^9,10^. For brain MRI traits, we analyzed a different set of regional brain volumes and DTI parameters compared to previous ExWAS^16,17^. Additionally, we discovered associations between rare variants and fMRI traits, which had not been previously reported.

Variant-level tests and gene-based burden tests provided different sets of associated genes. Although it has been widely accepted that gene-based tests enhance signal detection^16,17^, annotating the deleterious variants is still an active research area. In group-based tests, variants are collapsed into functional-frequency groups, but different approaches may lead to inconsistent and/or incomplete results^127^. Therefore, it is helpful to leverage the annotation information from diverse resources to better empower the set-based association test. We incorporated AlphaMissense^26^ as part of our annotations, resulting in the discovery of an additional set of genes (*GOLM1*, *KCNA5, LPAR3, PIGX, TAGLN,* and *WIPF3*). AlphaMissense is a novel deep learning model for pathogenic missense variant prediction and achieved good performance in predicting the unknown clinical significance of many missense variants.

Over half of the identified rare variant signals were within 1Mb of GWAS common variants. These rare variant associations could prioritize the causal genes among many identified GWAS loci and uncover novel signals. Human organ MRI traits are widely used as endophenotypes for diseases and health-related traits; our rare variant results deepen our understanding from two aspects. First, we showed that many of the identified gene-trait pairs were consistent with existing literature on the associated genes and their links to organ-related diseases or complex traits. This alignment includes findings from previous GWAS and ExWAS, observed clinical outcomes related to these genes, and animal models that elucidate the biological mechanisms connecting the gene to the phenotype. Second, we observed a significant enrichment of potential drug targets among our rare variant signals. This suggests that rare variant imaging genetics can play a role in identifying and repurposing drug targets, as well as understanding potential side effects during drug development. These findings have practical implications for both real-world clinical applications and future scientific research.

Along with these significant new insights, the present study has a few limitations. A primary limitation is the relatively modest sample size for identifying rare variant associations with MRI traits. For example, when the sample size increased from an average of *n* = 30,739 in the phases 1 to 3 analysis to an average of *n* = 40,038 in the phases 1 to 5 analysis, we observed a substantial increase in identified signals (84 vs. 174, that is 59 variant-level associations and 25 unique gene-trait pairs in the phases 1 to 3 analysis, and 107 variant-level associations and 67 unique gene-trait pairs in the phases 1 to 5 analysis). It is reasonable to hypothesize that a large number of rare variant associations have not been discovered for many MRI traits. Compared with recent ExWAS studies for other phenotypes encompassing nearly or more than 500,000 participants^16,17,21,62,128,129^, there remains substantial room for improvement in imaging studies. This is especially important for MRI traits that have a highly polygenic architecture, such as regional brain volumes, and is also critical for simultaneously comparing multiple traits. As the UKB imaging project completes data collection from 100,000 subjects^130^, we anticipate detecting additional signals and obtaining more robust results. This will not only help identify associated rare coding variants and genes but also elucidate the genetic architecture, such as burden heritability and burden genetic correlation, facilitating downstream analyses for rare coding variants.

Another limitation is the lack of diversity in imaging ExWAS data resources. First, no independent non-UKB database currently exists that combines both MRI traits and WES data in a sample size comparable to the UKB study. As discussed above, rare variants typically require a large sample size for detection and replication. Although our study leveraged the largest available dataset and had enhanced statistical power, we were unable to perform independent non-UKB replications because many rare variants may not be observed in smaller samples. Consequently, we used an internal validation procedure to assess the robustness of our findings. Nonetheless, future independent studies with more diverse data resources are essential to replicate the signals we identified. Second, our analysis primarily focused on individuals of European ancestry. It is critical to extend these studies to underrepresented ancestry groups in genetic research as data become available. Different ethnicities may exhibit heterogeneous genetic architectures, and including a diverse range of ancestries could provide a more comprehensive understanding of genetic influences on organ structure and function.

In conclusion, we have identified associations between rare coding variants and imaging traits across human organs. These findings enhance our understanding of the genomic mechanisms that regulate organ structure and function, potentially contributing to the identification and prioritization of novel targets for pre-clinical and clinical drug development. Looking ahead, it is important to increase sample sizes, integrate data from diverse populations, and expand ExWAS to include a broader spectrum of imaging phenotypes. We anticipate that future collaborative efforts will further elucidate the genetic landscape of human organs, advancing our knowledge of human biology and health.

## METHODS

Methods are available in the ***Methods*** section.

## Data Availability

The individual-level data used in this study can be obtained from UK Biobank (https://www.ukbiobank.ac.uk/). Other datasets in this paper include: Genebass (https://app.genebass.org/), GWAS summary statistics for imaging traits in BIG-KP (https://bigkp.org/), dbNSFP v.4.5a (https://sites.google.com/site/jpopgen/dbNSFP), the Therapeutic Target Database (https://idrblab.net/ttd/), and the GTEx dataset v8 (https://gtexportal.org/home/). The full ExWAS summary statistics generated in this study will be deposited in Zenodo upon publication.

## ACKNOWLEDGEMENTS

Research reported in this publication was supported by the National Institute On Aging of the National Institutes of Health under Award Number RF1AG082938 (B.Z. and H.Z.). The content is solely the responsibility of the authors and does not necessarily represent the official views of the National Institutes of Health. The study has also been partially supported by funding from the Purdue University Statistics Department, Department of Statistics and Data Science at the University of Pennsylvania, Wharton Dean’s Research Fund, Analytics at Wharton, and Perelman School of Medicine CCEB Innovation Center Grant (B.Z.). The study has also been partially supported by the National Institute On Aging of the National Institutes of Health under Award Numbers U01AG079847 and R01AR082684 (H.Z.). This research has been conducted using the UK Biobank resource (application number 76139), subject to a data transfer agreement. We would like to thank the individuals who represented themselves in the UK Biobank for their participation and the research teams for their efforts in collecting, processing, and disseminating these datasets. We would like to thank the research computing groups at the University of North Carolina at Chapel Hill, Purdue University, and the Wharton School of the University of Pennsylvania for providing computational resources and support that have contributed to these research results.

## AUTHOR CONTRIBUTIONS

Y.F. and B.Z. designed the study. Y.F., Jie C., Z.F., D.Y.Z., and Z.S analyzed the data and generated the results. Tengfei L., S.H., Z. J, P.Y.G., X.Q., Ting L., and H.L processed the MRI data. Julio C., J.L.S., P.F.S., R.W., A.N., M.D.R., J.O., W.W., D.J.R., and H.Z. provided feedback on study design and helped results interpretations. Y.F. and B.Z. wrote the manuscript with feedback from all authors.

## METHODS

### Ethics

This study made use of the data from UK Biobank (UKB) involving approximately 500,000 participants aged from 40 to 69 when recruited between 2006 and 2010 (https://www.ukbiobank.ac.uk/). The UKB study received the ethics approval from the North West Multi-centre Research Ethics Committee (reference number: 11/NW/0382) with informed consent obtained by all the participants. The present study was under UKB application number 76139.

### Multi-organ imaging phenotypes

The information of imaging phenotypes can be found in previous studies^1-4,10^. Briefly, we studied 596 imaging traits encompassing brain MRI traits^1-4^, heart CMR traits^10^, and abdominal MRI traits. Three major modalities of brain MRI were included. First, we included 101 regional brain volume traits^1^ derived from structural MRI. Second, we used 110 tract-averaged parameters^2^ from DTI capturing the microstructure of brain white matter. Third, for fMRI traits, we involved 76 node amplitude traits and six global functional connectivity traits based on ICA^3^ as part of resting fMRI traits; we also included 180 (90+90) parcellation-based^4,131^ resting fMRI traits and task fMRI traits. Regarding CMR traits, we made use of 82 traits^10^ including regional and local measurements from cardiac chambers and the aorta. For abdominal MRI traits, we contained 41 imaging traits based on MRI data of the liver, kidneys, lungs, pancreas, spleen, and body muscle/fat composition. A full list of these 596 traits can be found in **Tables S1**.

### Internal replication and joint analysis design

Given the limited sample size (average *n* = 40,038 across imaging traits with non-missing data) and lack of independent data sources, we prioritized our signals through joint analysis^30^ and adopted an internal validation procedure to evaluate the robustness of our discoveries. Specifically, we first restricted our sample to include only participants from phases 1 to 3 of MRI release (average *n* = 30,739) and then excluded any individuals related to phases 1 to 3 participants in the rest of the sample from phases 4 to 5 of MRI release (average *n* = 8,989, relatives of the phases 1 to 3 were removed). The joint sample included all the individuals from phases 1 to 5 and was used to report significant results. To investigate the robustness of variant-level associations, we tested the associations on phases 1 to 3 individuals and phases 4 to 5 individuals separately. Then, focusing on the significant level and effect sign, we investigated (i) whether signals identified from the phases 1 to 3 sample could be replicated in the independent phases 4 to 5 sample and (ii) whether the signals identified from phases 1 to 3 sample had stronger evidence (i.e. smaller *P* values and concordant effect directions) in the joint sample. For gene-based burden tests, we noted that many rare mutations, especially those variants with MAF < 1 × 10^-4^, accounted for a large proportion of our discoveries but were not observed in the smaller independent dataset from phases 4 to 5 (average *n* = 8,989; even within a burden model with MAF cutoff at 0.01, the lack of those rare variants in the sample from phase 4 to phase 5 would make the burden associations incomparable). Thus, we only investigated whether the identified associations from phases 1 to 3 dataset had stronger evidence in the joint sample. Consequently, based on this internal replication procedure, we were able to expect the level to which our discoveries from the joint analysis could be replicated.

### Quality control for UK Biobank exome data

We used Plink v.2.0^132^ to conduct the quality control steps, restricting the sample to individuals with imaging data (*n* = 54,365). For exome sequencing data, we included all the variants that had a MAF below 0.01. Variant-level quality control excluded all the variants that had missing rate larger than 10% or had Hardy-Weinberg equilibrium *P* < 1 × 10^-15^. Sample-level quality control excluded any sample with missing rate over 10%. Consequently, no individual was excluded from the quality control process, while 8,127,841 variants remained for the downstream analyses before annotation.

### Functional annotation for protein-coding variants

We used Variant Effect Predictor^133^ (VEP v.108) for variant annotation. Each variant was mapped to the most severe consequence across the canonical transcripts. We defined the loss-of-function variants using the Loss-of-Function Transcript Effect Estimator^134^ (LOFTEE) plugin, which further collapsed stop-gained, essential splice, and frameshift variants into high-confidence predicted loss-of-function variants (hcpLoF or pLoF hereafter) or low-confidence predicted loss-of-function variants (lcpLoF). Missense variants were prioritized by dbNSFP^135,136^ (v.4.5a) plugin using five prediction algorithms^16^: SIFT^22^, PolyPhen2 HDIV^23^, PolyPhen2 HVAR^23^, LRT^24^, and MutationTaster^25^. Missense variants were defined as “int5 damaging missense variants” if predicted damaging or possible damaging by the intersection of all the five algorithms and “int1 damaging missense variants” if predicted damaging or possible damaging by any one of the five algorithms. Parallelly, we also used AlphaMissense^26^ to assign the missense variants to be “Alpha damaging” if predicted as “pathogenic”. Synonymous variants served as empirical null control to support the study-wise *P*-value threshold for burden tests. Predicted loss-of-function variants (including both hcpLoF and lcpLoF) and missense variants were included in our downstream association tests. They were referred to as non-synonymous variants or coding variants of interest (*n* = 2,143,707).

### Exome-wide association tests

The covariate adjustment for brain MRI, DTI parameters, brain fMRI, and heart CMR traits was consistent with our previous GWAS^1-3,10,119^. In short, we adjusted a set of basic covariates for all the imaging traits including age (at imaging), age-squared, sex, age-sex interaction, age-squared-sex interaction, imaging site, and the top 40 genetic PCs (for the phases 4 to 5 sample replication analysis, we only adjusted for top 10 genetic PCs). For brain structural MRI traits, we additionally adjusted for total brain volume (for traits other than itself). For brain fMRI traits, we additionally adjusted for the effects of volumetric scaling, head motion, head motion-squared, brain position, and brain position-squared. For heart CMR traits, we additionally adjusted for the effects of standing height and weight. For abdominal MRI traits^137^, we additionally adjusted for the effects of standing height and body mass index.

We used REGENIE (v. 3.1.3)^138^ to conduct the exome-wide association tests. Common variants from genotyping array data (MAF > 0.01, genotype missing rate < 10%, Hardy-Weinberg equilibrium *P* > 1 × 10^-15^, LD pruning with r^2^ < 0.9) were included in the step 1 of REGENIE to capture genome-wide polygenic effects. Then, the predictors obtained from step 1 were used in step 2 for both variant-level tests and gene-based burden tests. For variant-level tests, we tested all the coding variants of interest as defined above. For gene-based burden tests, we included three general types of burden masks: (i) hcpLoF variants only, (ii) damaging missense variants only, and (iii) the combinations hcpLoF, lcpLoF, and damaging missense variants. For damaging missense variants, we further defined three categories, that is “int5 damaging”, “int1 damaging”, and “Alpha damaging” missense variants as described above, which resulted in seven finer variant sets for each gene: (i) hcpLoF variants only, (ii) “Alpha damaging” missense variants only, (iii) “int5 damaging” missense variants only, (iv) “int1 damaging” missense variants only, (v) hcpLoF variants and “Alpha damaging” missense variants, (vi) hcpLoF variants and “int5 damaging” missense variants, and (vii) hcpLoF variants, lcpLoF variants, and “int1 missense” variants. Additionally, for each gene set, we considered four levels of variants based on the alternative allele frequency (REGENIE used alternative allele frequency for separation, but we would keep the notation MAF as they were mostly concordant in our analysis): singletons only, MAF ≤ 1 × 10^-4^, MAF ≤ 1 × 10^-3^, and MAF ≤ 1 × 10^-2^. We noted that some sets may not be testable due to lack of qualified variants (for example, considering a gene that only had variants with MAF > 1 × 10^-3^) while some sets would produce repetitive results (for example, considering a gene that only had variants with MAF ≤ 1 × 10^-4^). To account for these issues, we adopted an empirical null based *P*-value threshold for multiple testing adjustment as described below.

### Study-wise significance level for association tests

For variant-level associations, we applied the Bonferroni correction at 0.05 level, which yields a conservative *P*-value cutoff at 2.8 × 10^-10^ (= 0.05/178,280,016, that is 0.05 adjusted for all the variant-level tests for coding variants of interest, see Table S21).

Moreover, we also provided the results for the variant-level associations that passed the rare variant genome-wide significance threshold at 1 × 10^-8^ as suggestive evidence^139,140^. The *P*-value cutoff for phases 1 to 3 sample was 3.5 × 10^-10^ (= 0.05/142,935,657, Table S21).

For gene-based burden tests, Bonferroni correction for all the tests conducted may be too strict and inappropriate here since distinct burden models and MAF cutoffs were highly correlated and may produce repetitive results. Moreover, the dependence structure made it unclear whether the Benjamini and Hochberg procedure could provide valid false discovery rate control. Alternatively, we used resampling-based methods to derive the empirical distributions, which suggested setting 1 × 10^-9^ as our conservative *P*-value threshold for burden tests. Specifically, we investigated the empirical null distributions of *P*-values from both permutation test^61,62^ and synonymous burden test^62,141^ separately. We conducted the permutation for the imaging traits once for every phenotype while preserving the genetic structure and the burden models^61,62^. At the tail of 243,691,511 *P*-values derived from the permutation-based null test, we only observed 5 results that had *P*-values smaller than 1 × 10^-9^ (Table S22). Therefore, based on this permutation-derived threshold at 1 × 10^-9^, the expected false discoveries would be 5 out of 224 significant associations (3 out of 67 significant gene-trait pairs) across all burden models and imaging phenotypes.

Furthermore, we found that under this permutation-based *P*-value cutoff, only synonymous variants in *NOSTRIN* and *HIGD1B* were observed to be associated with the mean diffusivity of the cingulum (cingulate gyrus) tract in brain white matter and the volume of left rostral middle frontal region, respectively, at the tail of the distribution of 40,545,911 *P*-values from synonymous burden tests (Table S23). These two genes were previously reported to be associated with white matter^142,143^ and other brain-related traits^144,145^, although the biological significance of the roles of synonymous variants remains unclear. Nevertheless, the very few significant associations further supported the validity of our choice of this *P*-value cutoff through a complementary perspective as synonymous variants generally would not contribute to the gene-trait associations^62,141^. We observed similar patterns of the tail distributions of *P*-values from permutation test and synonymous burden test from the phases 1 to 3 sample (Tables S24-S25). Thus, the *P*-value cutoff for burden tests of this sample (used in our internal replication) was set to be 1 × 10^-9^. We also provided a more liberal *P*-value cutoff at 1 × 10^-8^ as suggestive evidence (approximately empirical false discovery proportion less than 0.1, Tables S22-S23) for gene-based burden test.

### Cross-reference with previous GWAS

For the comparison with GWAS signals, we leveraged the summary statistics derived from previous studies^1-4,10^ on the same study cohort. The *P*-value cutoff for GWAS signals was set to be the Bonferroni adjustment for all the traits (*P* < 5 × 10^-8^ / trait numbers for each category) as described previously. Moreover, we focused on reported independent variants^146^ for GWAS signals and examined whether our exome-wide signals fell within the 1Mb range of such independent GWAS signals to prioritize the effector genes among thousands of identified GWAS loci. The genetic build for previous GWAS was GRCH37, so we did liftover^147^ for our exome-wide results back from GRCH38 to GRCH37 and made comparisons based on the same genetic coordinates.

### Imaging genetics and drug detection

We downloaded Therapeutic Target Database (TTD, last updated by January 10th, 2024)^120^ and evaluated all the entries documented in TTD that were claimed to be approved drug targets, in clinical trials, or supported by literature. We performed enrichment tests to see whether the identified genes in our gene-based burden tests were enriched in TTD drug target genes. We conducted Fisher’s exact tests for two classes of drug target genes in TTD. Specifically, we first included all drug targets that are either approved drug targets or in clinical trials (documented as ‘Successful’ or ‘Clinical trial’ in TTD), and then additionally included drug targets that were supported by literature (documented as ‘Literature-reported’ in TTD).

### Ultra-rare burden heritability and genetic correlation with health-related traits

We used the recently proposed BHR^27^ method to quantify the burden heritability and the burden genetic correlation for rare variants. BHR required one to divide the rare variants into multiple subgroups based on MAF cutoff and annotation outcome. Following the practical guide and previous examples, we investigated the genetic architecture of ultra-rare variants (MAF < 1 × 10^-4^) in our sample across 596 imaging traits. We used “univariate” mode and ran BHR to estimate the burden heritability for ultra-rare pLoF variants and “int1 damaging” missense variants. Moreover, we downloaded 186 health-related traits or diseases from Genebass^17^ to estimate the burden genetic correlation with imaging traits in terms of the ultra-rare pLoF variants. Among the 596 imaging traits examined, those that showed significant heritability after multiple testing adjustment (*P* = 8.4 × 10^-5^ = 0.05/596, that is Bonferroni adjusted for 596 imaging traits) were included in burden correlation analysis. Then, we ran “bivariate” mode of BHR to calculate the burden genetic correlation for the selected imaging traits and downloaded phenotypes.

## Code availability

We made use of publicly available software and tools. The code used in this study will be deposited in Zenodo upon publication.

---

**Table 1.** Significant gene-trait pairs for exome-wide gene-based burden test. Here we present non-redundant results for gene-trait pairs that passed the stringent threshold at *P* < 1 × 10^-9^. As a gene-trait pair may appear in multiple burden models and MAF cutoffs, only the association with the smallest *P*-value is included. Column “Phenotype_ID” uniquely defines a phenotype while column “Phenotype_info” provides additional information of the specific phenotype if applicable. Specifically, for abdominal MRI traits, the UKB data category is included in “Phenotype_info”; for CMR traits and DTI parameters, the full name is included in “Phenotype_info”; for all the fMRI traits (including ICA-based and parcellation-based ones), the network name is included in “Phenotype_info”. In the column “Organ/Category”, we further separate non-brain traits (i.e. abdominal MRI and CMR traits) to the corresponding organs. More details about the phenotype information can be found in **Table S1**, and a more comprehensive version of this table can be found in **Table S6**.

